# Loss of Y is associated with multi-omic changes in immune cells from Alzheimer’s disease patients

**DOI:** 10.1101/2023.02.07.23285520

**Authors:** Marcin Jąkalski, Edyta Rychlicka-Buniowska, Hanna Davies, Daniil Sarkisyan, Maciej Siedlar, Jaroslaw Baran, Kazimierz Węglarczyk, Janusz Jaszczynski, Janusz Ryś, Vilmantas Gedraitis, Alicja Klich-Rączka, Lena Kilander, Martin Ingelsson, Jan P. Dumanski, Jakub Mieczkowski

## Abstract

Alzheimer’s disease (AD) is a common and increasing societal problem due to the extending human lifespan. In males, loss of Y (LOY) in leukocytes is more prevalent in AD patients. We studied DNA methylation, gene expression and other epigenetic changes in monocytes and granulocytes with and without LOY from male AD patients and controls. New candidate genes were identified and numerous genes already implicated in AD pathogenesis were confirmed. We show that multi-omics of leukocytes can define AD candidate genes and we strengthen the role of LOY in disease development. The LOY-associated differences in DNA methylation levels were predominantly observed in regulatory regions, and supported by expression analysis showing down-regulation of immune genes. The single-cell transcriptomics highlighted that the AD patient-specific immune activation dominates over LOY-specific activation. Our findings agree with the hypothesis that age-related dysfunction of immune cells contribute to AD and that LOY is reflected by higher-level epigenetic changes with an AD-specific pattern.

## INTRODUCTION

Loss of chromosome Y (LOY) in leukocytes from aging males is the most common post-zygotic mutation, detectable in whole blood DNA from >40% of men above the age of 70 years (Thompson et al. 2019), reaching 57% in the analysis of 93-year-old men (Forsberg et al. 2019). Recent single-cell analyses of 29 aging men (median age 80 years) identified cells with LOY in every studied subject (Dumanski et al. 2021). LOY has also been observed in other tissues although with lower frequencies (Forsberg et al. 2019; Haitjema et al. 2017) and serial analysis of blood samples showed that LOY is a dynamic process (Danielsson et al. 2019; Bruhn-Olszewska et al. 2022). Major risk factors for LOY include age, smoking and germline predisposition (Thompson et al. 2019; Forsberg et al. 2019; Dumanski et al. 2016, 2015). LOY causes a clonal expansion of affected cells and may coexist with CHIP mutations (Ljungström et al. 2021). Recent analysis also suggests that LOY affects different lineages of hematopoietic cells with different frequencies and that it could play a role in dysregulation of autosomal genes through LOY-Associated Transcriptional Effects (LATE) in a pleiotropic manner (Dumanski et al. 2021). Moreover, dysregulation of immune genes was pronounced in LOY cells (Dumanski et al. 2021; Mattisson et al. 2021).

LOY has been associated with increased risk for all-cause mortality as well as with chronic and acute age-related diseases inside and outside of the hematopoietic system (Haitjema et al. 2017; Forsberg et al. 2017; Dumanski et al. 2016; Sano et al. 2022; Bruhn-Olszewska et al. 2022; Loftfield et al. 2018, 2019). Notably, LOY showed strong association with late-onset Alzheimer’s disease (LOAD), as males with LOY had 6.8-fold greater risk for Alzheimer’s disease (AD) diagnosis (Dumanski et al. 2016). This could be compared with the strongest LOAD genetic risk factor, the _ε_4 allele of the apolipoprotein E gene (*APOE*). The presence of one or two copies of *APOE* _ε_4 increased the risk to develop LOAD by a factor 3 up to 15-fold in a dose-dependent manner (Cacace et al. 2016; Guo 2021). The replication of the association between LOY and AD in additional cohorts, also applying a different methodological approach, has recently been shown in two reports (García-González et al. 2022; Palmer et al. 2022). Other investigations of transcriptome further suggested that LOY might indeed be important in the pathogenesis of AD (Caceres et al. 2020). Moreover, recent study of LOY in human brains from healthy aging subjects has shown that microglia had the highest number of LOY cells, and that there was a significant increase of LOY in microglia from male AD donors, which suggests a potential functional role of LOY in the pathogenesis of AD (Vermeulen et al. 2022). The process of clearance of amyloid plaques by microglia in the brain and clearance of circulating amyloid-beta from blood by monocytes has further been suggested as an important disease mechanism in AD (Yeh et al. 2016; Chen et al. 2020). Brain microglia and circulating monocytes represent functionally closely related cells, and monocytes from blood can migrate across the blood-brain barrier in response to inflammatory stimuli in various diseases, including AD (Fiala et al. 2005; Feng et al. 2011; Thériault et al. 2015; Zhao et al. 2020).

AD is the most common form of dementia and already a major public health problem, which is expected to worsen further due to the ever-increasing human lifespan (Prince et al. 2013). Depending on whether onset of symptoms occurs before or after 65 years of age, AD can be broadly defined into LOAD (∼90% of AD patients) and early onset Alzheimer’s disease (EOAD) (Cacace et al. 2016). Whereas EOAD sometimes can be explained by mutations in either of three genes (*APP, PSEN1, PSEN2*), LOAD is believed to be caused by complex genetics in combination with environmental factors (Jansen et al. 2019). For LOAD, analyses of twins have suggested a high heritability of AD with estimates ranging between 60-80% (Gatz et al. 2006). Moreover, in addition to *APOE* genome wide association studies (GWAS) have identified many AD risk genes (Cacace et al. 2016; Lambert et al. 2013; Jansen et al. 2019; Bellenguez et al. 2022).

Multiple studies have shown that epigenetic processes are often dysregulated and might play a role in development and progression of AD. DNA methylation at CpG sites provides a stable epigenetic modification that usually silences (alternatively promotes in case of demethylation) the transcription of adjacent gene(s) (Moore et al. 2012; Hughes et al. 2020). Altered DNA methylation levels in whole blood DNA were identified as associated with worse cognitive performance and accelerated rate of AD progression (Li et al. 2021; Chouliaras et al. 2018; Madrid et al. 2018; Roubroeks et al. 2020). Furthermore, covalent modifications of histones, and higher-level three-dimensional structures (such as topologically associating domains, TADs), are other examples of dynamic epigenetic regulators. TADs constitute one of the forms of chromosomal organization within a nucleus into functional compartments (Tena & Santos-Pereira 2021). Genes bound by the same TAD are usually regulated in a coordinated manner, as TADs facilitate interactions between the genes and their distant regulatory elements. DNA methylation and histone modifications, such as histone methylation and acetylation, are in a constant interplay (Rose & Klose 2014; Miller & Grant 2013). Recent studies showed that epigenetic processes are often dysregulated in AD patients and that the epigenetic control of enhancers thus may have an important pathogenetic role (Li et al. 2019; Winick-Ng & Rylett 2018).

We have taken here a novel approach to study AD by incorporating multi-omics data (DNA methylation, gene expression) and the LOY status of the studied individuals, in an attempt to delineate candidate genes that might be involved in the pathogenesis. Similarly, we explored epigenetic changes in the intergenic regions. We took advantage of pure flow cytometry-sorted populations of granulocytes and monocytes, with LOY or in normal state. We hypothesized that LOY could be associated with global changes of DNA methylation and could lead to characterization of specific candidate genes, as well as trigger other epigenetic changes in AD, i.e., histone modifications and 3D structure of DNA.

## RESULTS

### Study subjects and measurements of LOY

We collected blood samples from 73 male individuals (median age 74 years, age range 61-90 years), which encompassed 43 AD patients with and 30 healthy controls, without a prior diagnosis of AD or cancer (Supplementary File 1 - Table S1). The peripheral blood leukocytes were sorted using fluorescence-activated cell sorting (FACS) to obtain pure cell populations of granulocytes and monocytes. The samples studied here were chosen out of subjects reported previously (Dumanski et al. 2021) and the only selection criterion was the availability of sufficient amount of DNA, to perform CpG methylation profiling on EPIC beadchip.

For each cell type, we estimated the levels of LOY mosaicism using SNP arrays, as described previously (Forsberg et al. 2014; Dumanski et al. 2016). The obtained mLRRY values (median Log R Ratio of probes located in the male specific part of chromosome Y) were transformed to estimate the percentage of cells with LOY (%LOY) in each sample (Danielsson et al. 2019). We could distinguish samples with high levels of LOY, and these were more frequent among AD patients as compared to controls (Figures 1A and 1B). Dependent on the estimated %LOY, samples were further divided into “*LOY”* and “*non-LOY”* groups (34 and 72 samples, respectively, Supplementary File 1 - Table S1). We used a 30% cutoff, *i*.*e*., where 30% of the cells have LOY, as presented previously (Dumanski et al. 2016; Danielsson et al. 2019). The AD samples presented a clear bimodal distribution (Figure 1B), and the 30% threshold separated the two peaks of data. Matched samples from granulocytes and monocytes (collected from same patients) presented a high concordance of the LOY estimates (Pearson correlation = 0.97, Figure 1C). Among the unmatched samples (left and bottom panels on Figure 1C) the presence of LOY was also associated with AD (9 out of 12).

**Figure 1.**
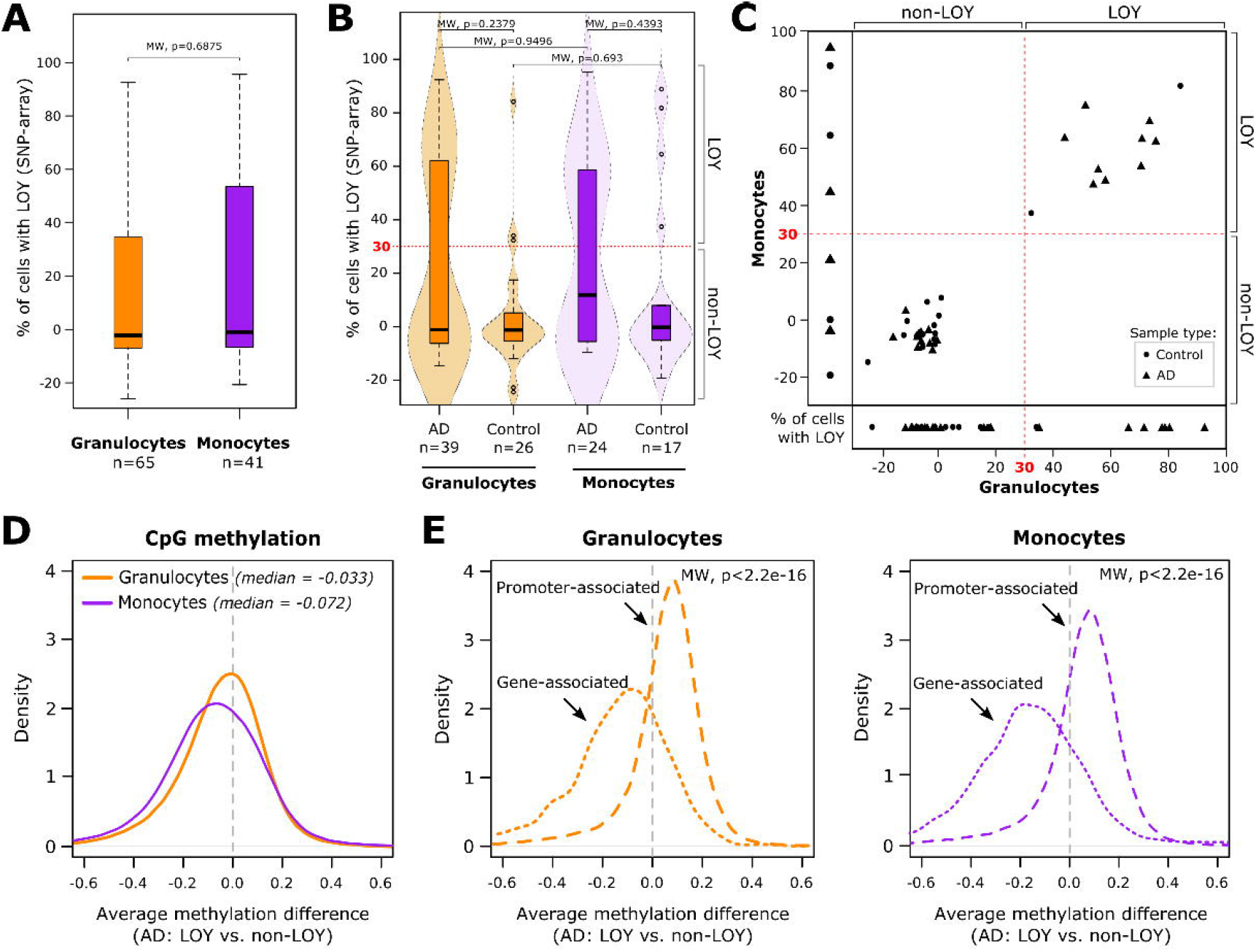

### Genome-wide profiling of DNA methylation landscape

Using the Infinium MethylationEPIC Beadchip we performed genome-wide DNA methylation analyses. Specifically, we obtained methylation data for 65 granulocyte (39 ADs, 26 controls) and 41 monocyte (24 ADs, 17 controls) samples covering 835,424 CpG sites after dropping the problematic probes with known SNPs (LaBarre et al. 2019). Initial inspection of the distribution of differences in average methylation values (M-values) between the four studied groups and per all individual genomic loci showed moderate differences (median of average difference ranging from -0.07 to 0.02, Supplementary Figure S1). The largest difference in global DNA methylation was observed among the monocyte samples (Figure 1D). Here, the monocytes from AD patients not affected by LOY were clearly more methylated than the monocyte-*AD-LOY* samples (the difference in average M-value shifted more towards the negative values). In the corresponding granulocyte samples this difference was much smaller.

Delving deeper into the inter- or intragenic localization of CpG probes and their functional annotation, e.g. localization within genes’ regulatory elements, revealed pronounced differences. Loci overlapping promoters and CpG islands showed higher methylation values in the LOY samples (more positive values of average methylation differences), both in granulocytes and monocytes (Figure 1E, Supplementary Figure S1), and these differences were significant both when compared against all genomic sites and gene-associated sites only (*p* < 2.2e-16, Mann-Whitney test, MW).

### Identification of differentially methylated sites

To identify LOY-related DNA methylation changes, and specifically those distinct for AD, we performed a differential methylation analysis. We compared the *LOY* against the *non-LOY* groups within the cohorts of AD patients and controls, as well the AD patients against controls between the matched *LOY*/*non-LOY* groups. CpG probes located on the Y chromosome were excluded from the analyses (see Methods). In total, based on these comparisons we identified 17,647 and 2,913 significantly differentially methylated probes (hereafter DMPs; adjusted p-value <0.05 and absolute M-value difference >0.5) in granulocytes and monocytes, respectively (Table 1, Supplementary File 1 – Tables S2-S7). Among the AD patients, hypomethylation in LOY samples dominated (Figure 2A), both in granulocytes (7,815 of 8,292 DMPs, 94%) and monocytes (1,578 of 1,657 DMPs, 95%). Additionally, out of the above DMPs 2,953 in granulocytes and 1,239 in monocytes were differentially methylated only between the AD LOY vs. AD non-LOY samples (Supplementary File 1 – Tables S8–S9). This suggests an AD-specific effect of LOY on DNA methylation. In granulocytes, but not in monocytes, the magnitude of hypomethylation increased significantly among these AD specific DMPs (*p* < 2.2e-16, MW test) (Figure 2A, bottom panels). Investigation of the genomic distribution of the identified DMPs showed that they were mainly localized within the gene body, intergenic regions, and the so-called open sea regions (see Supplementary Text). Finally, a comparison between granulocytes and monocytes showed that they shared 886 DMPs, all with a preserved direction of methylation change (Figure 2B). Sites identified to have a significant change in DNA methylation only in AD samples will be the main focus of further description. Additionally, to corroborate the DMP analyses, we also identified differentially methylated regions (DMRs, see Supplementary Text).

**Figure 2.**
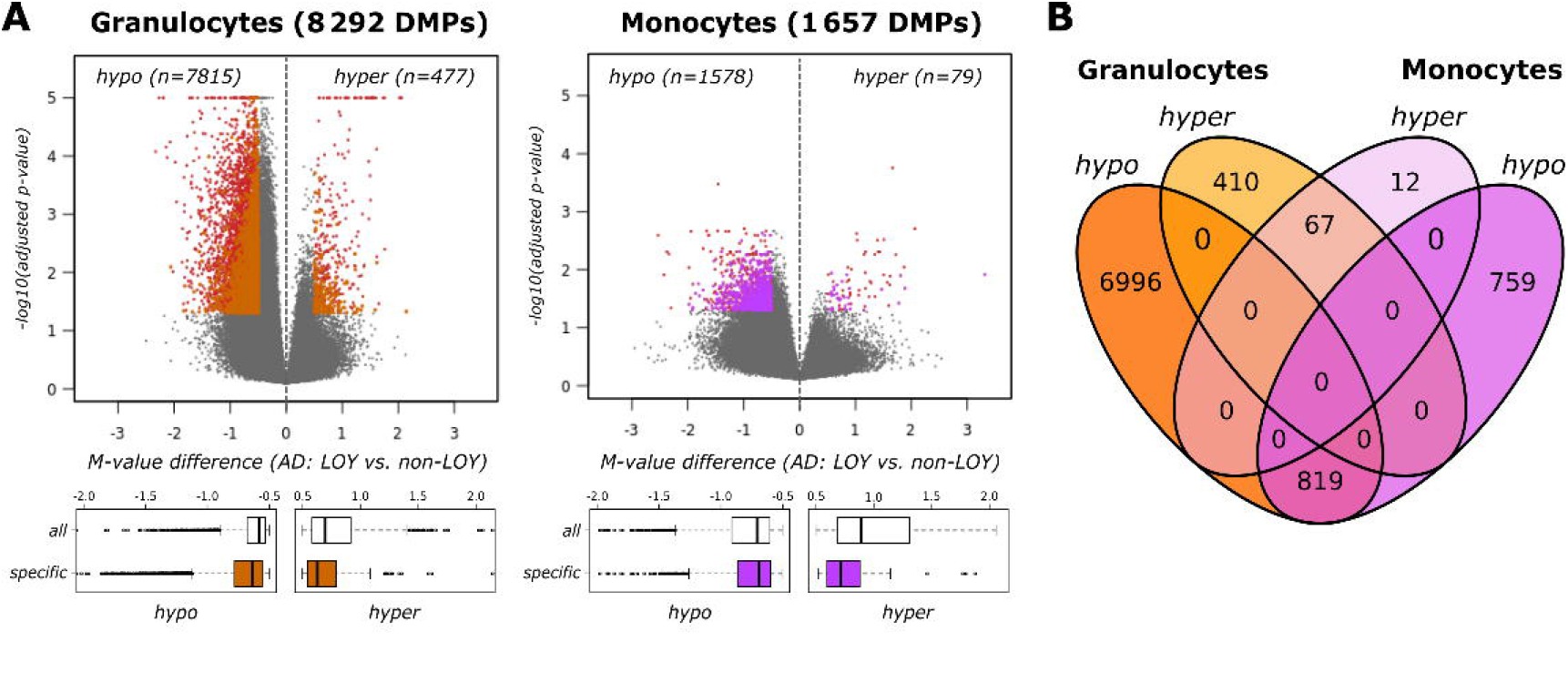

### Gene-associated methylation changes

We next analyzed genes associated with the identified DMPs (hereafter DMGs, Supplementary Figure S3A and B, Supplementary File 1 – Tables S8-S9) between AD *LOY* and AD *non-LOY* groups. In total, we found 2,022 such genes in granulocytes and importantly 1,045 of these genes had at least one DMP within their promoters (+/-2 kb from transcription start site, TSS). The corresponding numbers for monocytes were 884 and 437 DMGs. A comparison of the two cell types showed that 301 DMGs were found both in granulocytes and monocytes (Supplementary Figure S3C). Many of the DMGs (∼66% in granulocytes and ∼45% in monocytes) were associated with the AD according to the OpenTargets (Ochoa et al. 2021) and GeneCards (Safran et al. 2021) databases (Supplementary File 1, Table S16 and S17). Additionally, 45 and 19 DMGs were previously found to exhibit a LOY-associated dysregulation in granulocytes and monocytes, respectively (Dumanski et al. 2021) (Supplementary Figure S3D).

### CpG methylation is linked with changes of gene expression

To further explore the effects of DNA methylation changes related to LOY in AD, we performed a transcriptomic analysis using bulk RNA-sequencing (RNA-seq). The analyses were conducted on available RNA from granulocyte and monocyte samples (31 individuals, see methods). We identified 1,153 and 1,180 genes demonstrating significant differential expression (DEGs) in granulocytes and monocytes, respectively (Figure 3A, Table 7, Supplementary File 1 – Table S10-S11). While our differential methylation analyses showed that hypomethylated DMPs dominated in AD samples, here the LOY-associated effect on gene expression in AD was less pronounced - there were 1.1 to 1.3 times more downregulated than upregulated DEGs in granulocytes and monocytes, respectively.

**Figure 3.**
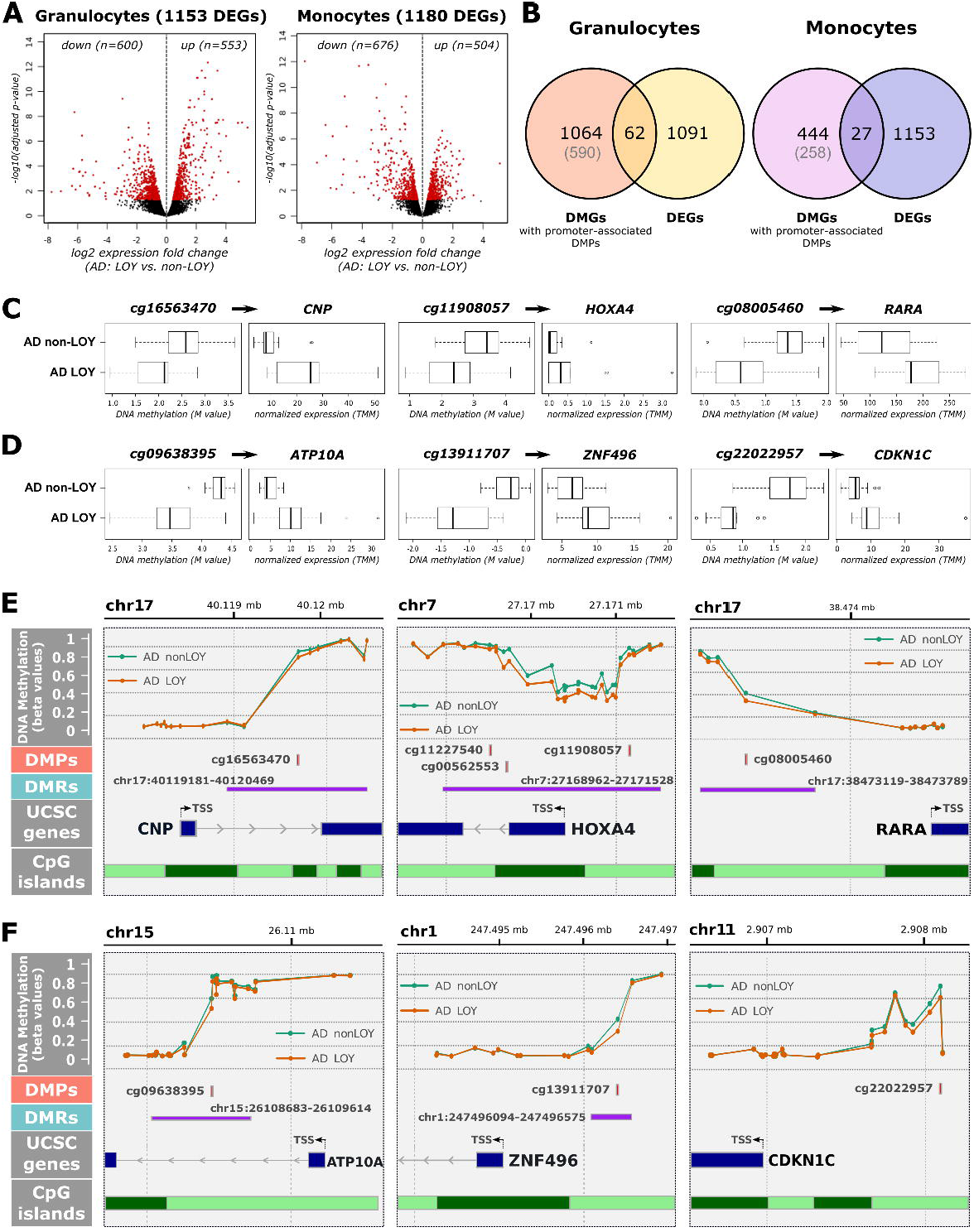

We further assessed to what extent the DMGs and DEGs overlapped. In granulocytes 100 genes were shared between the two analyses, and 62 of these had at least one LOY-associated DMP located within the promoter (Figure 3B, Supplementary File 1 – Table S12-S13). The corresponding numbers for monocytes were 51 and 27 (Figure 3B). Among granulocytes, DNA methylation of 29 of 66 (44%) probes was negatively correlated (Pearson correlation) with the expression level of the associated gene (Supplementary File 1 – Table S14), which is in line with the canonical model of regulation by CpG methylation (Moore et al. 2012). Similarly, 27 DEGs in monocytes had differentially methylated sites within their promoters and methylation changes of 16 of 31 probes (51%) were negatively correlated with expression of the associated gene (Supplementary File 1 – Table S14). Importantly, more than 50% of DMGs had no or very little measured expression in the bulk RNA-seq data hence were excluded prior to the DE analyses (Figure 2B, and methods).

### Examples of DMGs with LOY-associated transcriptional effect

The *CNP* gene (2’,3’-Cyclic Nucleotide 3’ Phosphodiesterase), which was significantly upregulated in the AD LOY group in granulocytes (log2 FC 1.24, FDR < 2.2e-16), had a single granulocyte-specific hypomethylated site (−0.61 M-value difference, FDR=0.032) located within its promoter (Figure 3C and 3E). The decreased gene expression of *CNP* was previously reported in AD brains (Natalia Silva et al. 2012). The *HOXA4* gene (Homeobox A4), which harbors over 30 CpG sites, had three hypomethylated probes identified, of which one (*cg11908057*, mean M-value difference -1.02, FDR=0.022) was present directly upstream of TSS (Figure 3C and 3E). *HOXA4* was also upregulated in LOY cells in granulocytes (log2 FC 2.21, FDR=0.013). Evidence for elevated DNA methylation levels spanning the *HOXA* gene cluster and association with AD were reported in brain samples (Smith et al. 2018; Altuna et al. 2019). The *RARA* gene (Retinoic Acid Receptor Alpha) was upregulated in LOY cells in granulocytes (log2 FC 0.6, FDR=0.03) and contained a single promoter-related DMP (average M difference -0.53, FDR=0.01) (Figure 3C and 3E). A decline in the transcriptomic levels of retinoic acid receptors was suggested to be involved in early stages of AD (Goodman & Pardee 2003; Khatib et al. 2020). Here the LOY cells had higher levels of *RARA* than the non-LOY ones.

*ATP10A* gene (ATPase Phospholipid Transporting 10A), significantly upregulated in LOY in monocytes (log2 FC 1.28, FDR=0) had a single hypomethylated site located within the promoter region (1st intron, mean M-value difference -0.78, FDR=0.03) (Figure 3D and 3F). Another example for monocytes included the *ZNF496* gene (Zinc Finger Protein 496). Cells with LOY had an increased expression of this gene in AD (log2 FC 0.74, FDR=0.016) and the single promoter-bound CpG site was significantly hypomethylated in LOY cells in monocytes (*cg13911707*, mean M-value difference -0.88, FDR=0.04). Similarly, the *CDKN1C* gene (Cyclin Dependent Kinase Inhibitor 1C) was upregulated in LOY cells (log2 FC 1.02, FDR= 0.003) and had a single DMP upstream of its TSS (average M-value difference -0.85, FDR=0.026). The latter three examples of genes at the intersection of DMGs and DEGs in monocytes were found to have an altered expression in the context of AD or LOY (Moradifard et al. 2018; Dumanski et al. 2021).

### LOY-associated epigenetic dysregulation beyond gene boundaries

The high number of DMGs and DEGs but a relatively limited overlap between these categories, led us to hypothesize that LOY may affect different modes of expression regulation. Many of the significant changes in DNA methylation that we identified cannot be linked directly to any specific gene (22-32% and 22-31% of DMPs in monocytes and granulocytes, respectively, Supplementary Figure S2A, Supplementary Text). They are located within the intergenic regions, and more than half of the sites lie outside of the CpG islands. Thus, to link LOY effect with the regulation of gene expression regulation we investigated the chromatin regions marked with the H3K4me3 and H3K27ac histone marks. Due to data availability (in the public domain), we performed this analysis only for monocytes (see Methods). We noted that the LOY-associated difference in methylation of active promoter regions (overlapping H3K4me3 and H3K27ac histone marks located near TSS) is skewed towards hypermethylation, as seen before for individual CpG probes. However, the difference between the AD and control samples, although significant (p <2.2e-16, MW test, Figure 6A) was not that smaller than for enhancers (regions of H3K27ac histone mark not overlapping H3K4me3 and far from TSS). In contrast, we observed a shift towards LOY-associated hypomethylation in AD, while the average methylation of enhancers peaked around zero for the controls. This suggested that effect of LOY on DNA methylation in larger in AD samples than in control samples.

The same pattern was observed when we investigated three-dimensional chromatin maps. We used the publicly available HiC data from the THP-1 human monocytic cell line from ENCODE Project to cap genes and our DNA methylation data under the TADs boundaries (Supplementary File 1, Table S15). Within TADs, comparison of LOY versus non-LOY methylation patterns (average methylation per group per TAD) showed that controls display only a slight skew towards the negative values (higher methylation in non-LOY) in average methylation difference. When analyzing AD samples only, the bias towards the negative values is much more pronounced and the distribution of these TAD-bound average differences is significantly distinct between controls and AD samples (p < 2.2e-16, MW test, Figure 6B). Altogether, these results suggest broader changes in gene expression regulation related to LOY.

### Regulatory gene networks further link DNA methylation and gene expression

The weighted gene co-expression network analysis (WGCNA) is used to identify clusters of genes with correlated expression patterns (Langfelder & Horvath 2008). This can help in determining gene modules that are significantly associated with LOY and AD. In granulocytes, we found 8 such modules, which showed differential expression between the investigated samples (Figure 4A, Supplementary File 1 Table SX). Most of these modules (6 out of 8) were upregulated in *LOY* (the individual genes that make up the modules are overall higher in *LOY* than in the *non-LOY* samples). The highest expression fold change was observed for the ‘light cyan’ module. In monocytes, there were 11 modules with significant change in gene expression related to LOY and 7 of these were upregulated. The module called ‘tan’ was the most up-regulated one.

**Figure 4.**
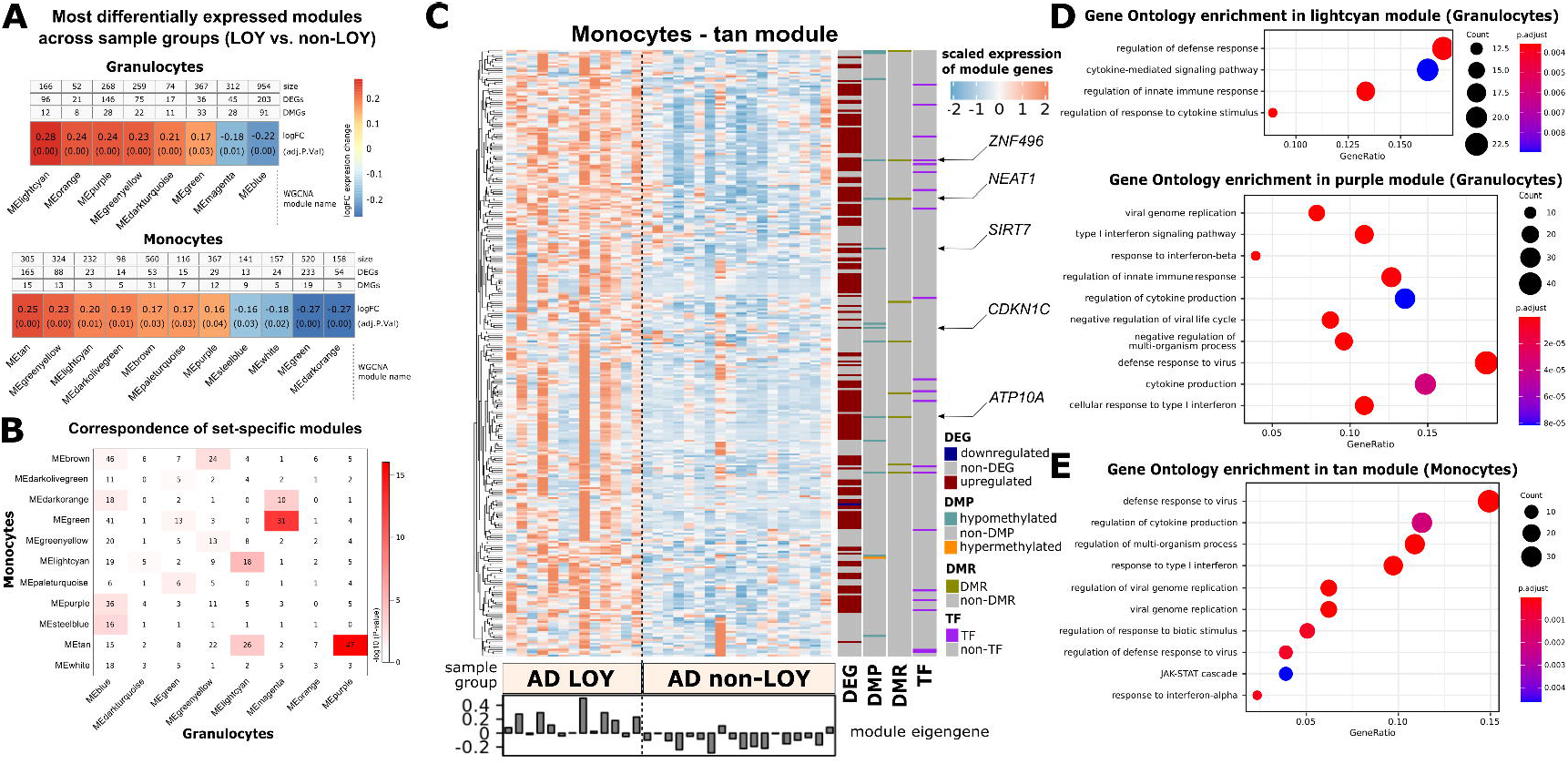

We next used a Fisher’s exact test to determine the significance of the correspondence between the two sets of modules computed for the two cell types (Figure 4B). The modules with the highest correspondence (set of overlapping module members) were ‘purple’ from granulocytes and ‘tan’ from monocytes. The latter also had a high correspondence to the ‘light cyan’ module from granulocytes. All these highly corresponding modules were among the most up-regulated in LOY and comprised 54-58% of DEGs and a number of DMGs (mostly hypomethylated) (Figure 4A and 4C). These three modules were significantly enriched in gene ontology terms related to immune processes (Figure 4D and 4E), which are known to be dysregulated in AD (Fani Maleki & Rivest 2019). Additionally, we found many transcription factors among the module members, several with dysregulated expression and a significant change in their DNA methylation status. The *ZNF496* transcription factor, together with four other genes at the intersection of DMG and DEG sets in monocytes belonged to the ‘tan’ module in monocytes (Figure 4C). Transcription factor binding sites (TFBS) of three and four transcription factors (members of ‘tan’) module were significantly enriched among LOY-associated DMPs and promoters of monocyte DEGs, respectively (FDR<0.05, Fisher’s exact test).

The downregulated modules in monocytes, e.g., ‘green’ and ‘dark orange’ also comprised many DMGs and DEGs, however they were not enriched in any functional categories. Still, among the module members we found multiple genes with functions related to regulation of transcription or apoptotic processes. It was shown that patients with AD are more prone to infections, which could be due to enhanced apoptotic death of their peripheral leukocytes (Bergman et al. 2002). The ‘blue’ module, which was significantly downregulated in granulocytes, was enriched in functions such as neutrophil degranulation and neutrophil activation (Supplementary Figure S5A). Neutrophil hyperactivation and accumulation is known to contribute to AD pathogenesis and cognitive impairment (Zenaro et al. 2015). Lastly, the ‘green’ module in granulocytes was enriched in gene ontology terms such as nucleosome assembly, chromatin assembly and DNA packaging (Supplementary Figure S5B). Dysregulation of proteins involved in genome organization, along with their altered localization, was confirmed to be associated with AD (Winick-Ng & Rylett 2018).

### LOY-related transcriptional effect revealed by single cell RNA-seq

To investigate the influence of LOY on cell subpopulations we applied single-cell RNA sequencing (10X Genomics). We analyzed cells collected from 26 AD patients (9 *LOY*, 17 *non-LOY*) and focused on monocytes only (see methods). Based on the presence of Y-linked expression we inferred the LOY status of cells and separated them into non-LOY (20,702) and LOY-cells (11,527) (Dumanski et al. 2021). Additionally, we divided the cells identified as monocytes into fine-grained labels, *i*.*e*., classical monocytes, intermediate monocytes, and non-classical monocytes (Monaco et al. 2019). The proportion of these cell subtypes differed significantly between the sample groups (Figure 5A).

**Figure 5.**
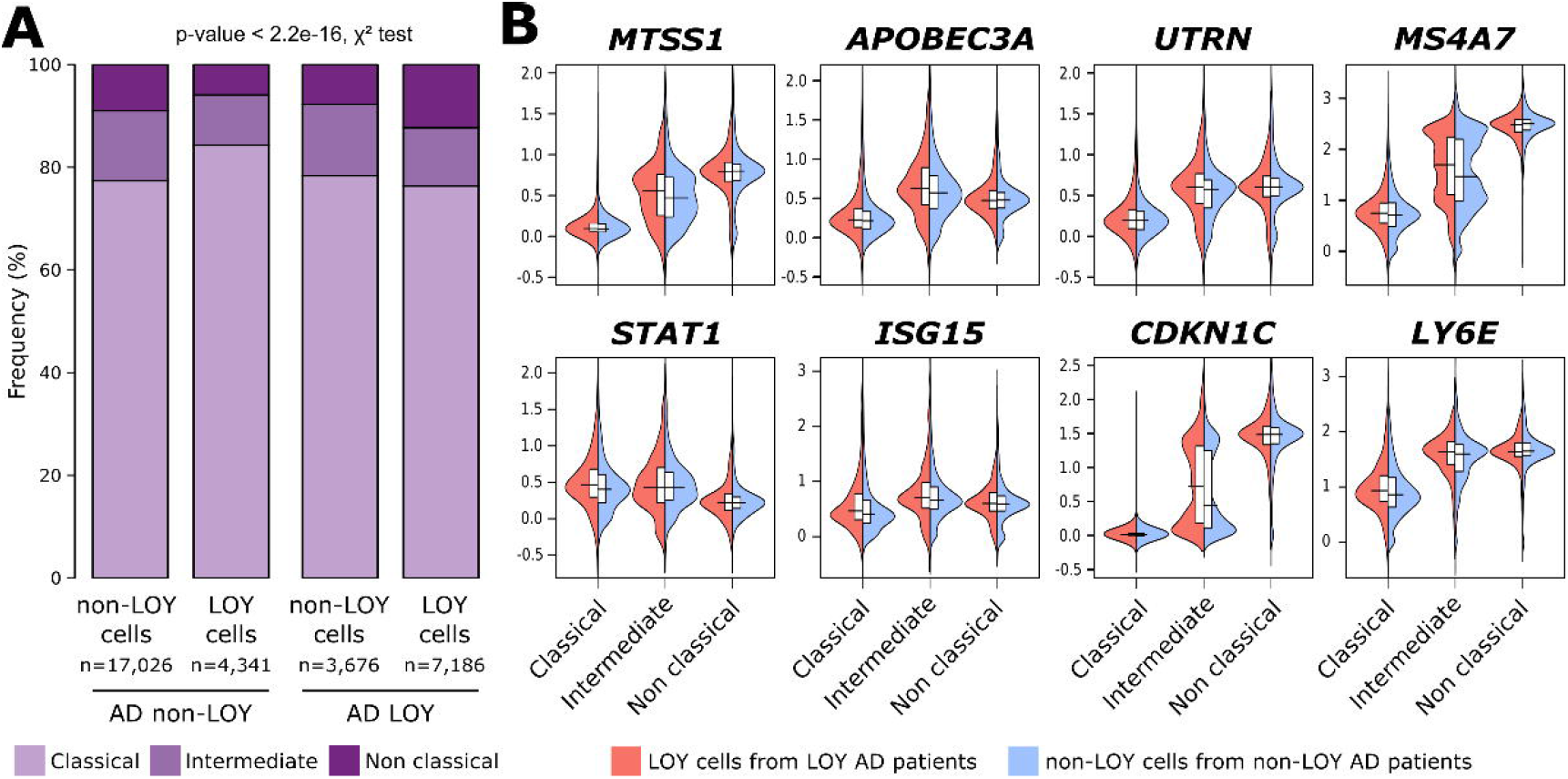

We next compared the expression in LOY cells from LOY AD patients against the non-LOY cells from non-LOY AD patients. Unlike in the analyses based on the bulk RNA-seq data (Figure 4A), here the upregulation in LOY was predominant (Supplementary File 1, Table X). In general, the total number of the scRNA-seq based DEGs (scDEGs) was much smaller, which most likely is related with the number of genes identified in all the samples. There were 101 scDEGs (74 up and 27 down) found for classical monocytes, and 101 (76 up, 25 down) for intermediate monocytes (for details see methods section). No statistically significant scDEGs were identified for non-classical monocytes. This highlights the subpopulation differences, especially compared with the obtained numbers of scDEGs where the subpopulations were not distinguished (68 up and 19 down genes). In total, all these comparisons yielded 148 differentially expressed genes (110 upregulated, 38 downregulated), of which 42 overlapped with DEGs identified with bulk RNA-seq from monocytes. Among the scDEGs upregulated in LOY, we again found significant enrichment of gene ontology terms related to immune/defense responses (Supplementary File 1, Table X). Moreover, scRNA-seq data revealed distinct expression patterns that depend on the monocyte subtype, and where the size of the transcriptomic effect of LOY was often cell type specific (Figure 5B).

When comparing the expression patterns of LOY cells vs. non-LOY cells within a group of patients, we could not identify as many differentially expressed genes (Supplementary File 1, Table X). Specifically, we found 30 DEGs by comparing monocytic LOY to non-LOY cells of AD non-LOY patients. We found another 41 genes by comparing LOY to non-LOY cells of AD LOY patients. Only ∼10% of scDEGs from the first single cell-based test overlapped with the DE genes obtained from these two within group comparisons (Supplementary File 1, Table X).

## DISCUSSION

We have studied sorted granulocytes and monocytes (with at least 95% purity) derived from male LOAD patients and healthy controls (without cancer and AD diagnoses in clinical history) for changes of CpG-methylation and differential expression analysis. These samples contained either high levels of mosaic LOY (>=30% of cells) or whose genome was predominantly not affected by LOY. From the AD and LOY point of view, the most important comparison was between the cells with or without LOY in AD patients. Furthermore, we have related the changes in the CpG methylation with RNA analysis from the same populations of cells, which allowed us to identify up-or down-regulated genes, presumably as a consequence of change in the methylation state.

According to the canonical model, hypomethylation within CpG islands and promoters should be associated with increased expression of the relevant gene and we identified many clear cases of pairs of differentially methylated genes (DMG) and differentially expressed genes (DEG) fitting into this model (Figure 3C-F, Supplementary File 1, Tables S12-S13). An interesting example of a gene with potential association to the progression of AD was the *HOXA4* gene with promoter hypomethylation and upregulation in granulocytes (Figure 3C and 3E). Li et al. has previously reported that hypomethylation in *HOXA4* was associated with a faster rate of cognitive decline (Li et al. 2021). Other genes from the HOX clusters, which are transcription factors and play diverse roles in development, were also found to be differentially methylated in AD (Smith et al. 2021). Another example following the canonical model is the CNP gene, encoding a very abundant protein in the central nervous system, which was promoter hypomethylated and significantly upregulated in LOY cells (Raasakka et al. 2015).

Our results showed that ∼50% of the identified DMGs had at least one promoter-associated differentially methylated probe (DMP). As mentioned above, the majority of the identified DMPs were hypomethylated. Hence, we should observe LOY-associated upregulation of the relevant genes. Our subsequent analyses of bulk RNA-seq data from corresponding patients showed, however, that only ∼50% of the DMGs were in fact expressed in both studied types of leukocytes (Figure 3B). Moreover, the numbers of upregulated and downregulated genes were similar, with slightly more of the downregulated ones, both in granulocytes and monocytes (Figure 3A). This could suggest that the LOY-associated effect of DNA methylation on gene expression is in fact often not following the canonical model of methylation changes and gene expression levels. The non-canonical associations between DNA methylation and gene expression (i.e., hypomethylation and downregulated expression, as well as hypermethylation with upregulated expression) has also been noticed in recent publications (Spainhour et al. 2019; Rauluseviciute et al. 2020; Anastasiadi et al. 2018). The *FOXK1* gene was an example of DMG and DEG, in both granulocytes and monocytes (Supplementary Figure S4). This gene harbored multiple CpG sites that were hypomethylated and located within the gene’s body. The expression of this gene, however, was downregulated in samples from AD patients belonging to the LOY group, thus not following the canonical model. Proteins from the *FOX* gene family can influence the development of neuronal precursors and maintenance of neurons (Genin et al. 2014; Maiese 2016). Moreover, *FOXK1* gene can serve as a transcription activator or repressor and it was found to be implicated in reducing virus replication, by promoting antiviral gene expression (Panda et al. 2015).

We also compared the genes that showed an overlap between DMGs and DEGs (Figure 3B) with the genes noted in the OpenTargets and GeneCards databases for AD. These lists are composed of 8,763 and 11,297 independent genes, respectively, with evidence in the literature or experimental data as potentially involved in AD. Each gene entry is given a score representing the strength of the association between the target gene and the disease, which could be used as a rough relative estimate of the importance of these genes for AD pathogenesis. We noticed many overlapping entries between DMGs and DEGs in monocytes and granulocytes and the OpenTargets and GeneCards (Supplementary Tables S16 and S17). The most prominent example was a dysregulation via hypomethylation of the amyloid precursor protein gene (*APP*) occurring only in granulocytes. However, these cells surprisingly showed a significant downregulation of *APP* transcripts. This finding should be followed by in depth analysis in other specific cell types, both from blood and brain.

Upon similar comparison between the OpenTargets/GeneCards and our results using only DEGs in monocytes and granulocytes, the lists with overlaps were considerably longer (Supplementary Tables S16 and S17). The most important example was the *TREM2* gene that was significantly downregulated in monocytes only. *TREM2* has attracted a considerable attention because of its role in microglia-mediated clearance of amyloid plaques and its binding to Apolipoprotein E (*APOE*). Moreover, it has been recently found to be a risk gene for AD (Ulland & Colonna 2018; Qin et al. 2021). Another interesting link between LOY, *TREM2* and DMGs/DEGs in monocytes and granulocytes are our results regarding dysregulation of multiple members of the *ADAM* gene family of disintegrin- and metalloproteases (Supplementary Tables S8-S11). One of the described functions of *ADAMs* is processing by cleavage of the *TREM2* ectodomain and its release as a soluble version (Yang et al. 2020).

CpG methylation status is known as highly cell-type specific (Zhou et al. 2017; Lokk et al. 2014). Hence, it was not surprising to find differences between monocytes and granulocytes, with the majority of the described DMPs not shared between these two cell types (Figure 2B). Furthermore, when considering the combined analysis of DMGs and/or DEGs and overlaps with OpenTargets/GeneCards databases, we should note that the RNA-seq is a relatively comprehensive method for assessment of expressed genes, compared to the sensitive but limited in numbers methylation analysis of CpGs performed on EPIC-arrays. It is currently estimated that the human genome contains about 28 million CpG sites (Babenko et al. 2017; Luo et al. 2014), while EPIC array covers around 850,000 CpGs, thus providing information for about 3% of CpGs.

At a single-cell level, loss of Y is a binary event removing the entire male-specific chromosome. This results in the disappearance of nearly 2% of the haploid genome, which should at least influence the packaging of DNA and chromosomes within a nucleus. Although poor in protein coding genes, chromosome Y contains many important genes with functions that go beyond sex determination. One such gene is *KDM5D*, which encodes a male-specific demethylase that targets H3K4me3 of histones (Arseneault et al. 2017). This chromatin landmark is usually found near TSSs and is an indicator of transcriptionally active genes. Thus, in the event of LOY, we could expect changes at the level of gene expression, triggered by chromatin remodeling. Our analyses of TADs and histone modifications showed an LOY-associated effect on DNA methylation within these regions (Figure 6). Additionally, we observed that LOY had an even more pronounced effect on DNA methylation in AD samples than in controls. This is in line with recent studies that showed dysregulation of epigenetic processes in AD and that regulation may occur through chromatin higher-order structures (Li et al. 2019; Winick-Ng & Rylett 2018; Kikuchi et al. 2019).

**Figure 6.**
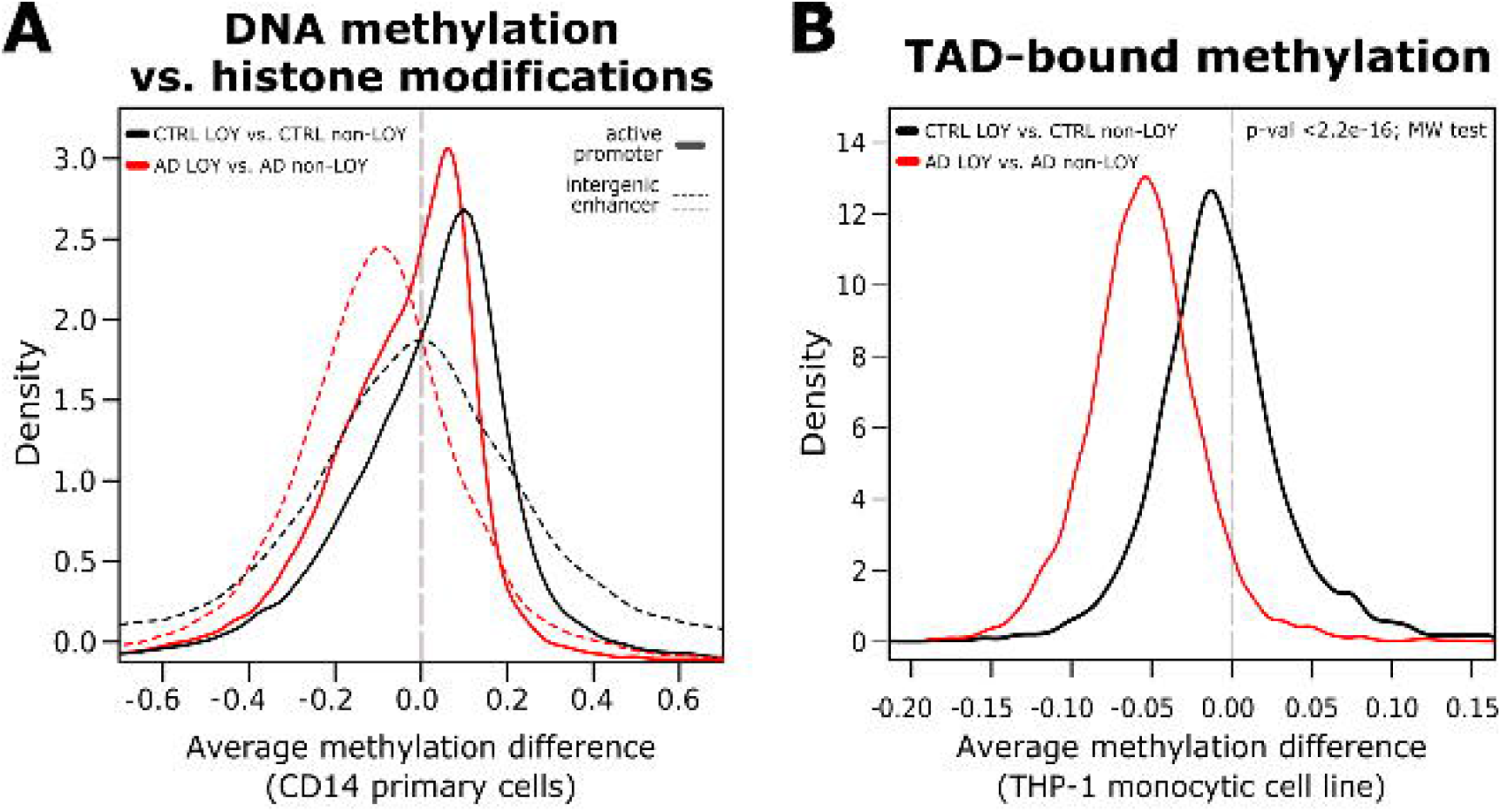

The observed changes in the DNA methylation profiles with regard to other chromatin properties shed new light on the gene expression regulation affected by LOY. We analyzed the profiles using publicly available data that describes the chromatin activation and three-dimensional chromatin structure. We showed significant differences in the regulatory regions, especially in regions remote from the promoters. This suggests that the loss of chromosome Y, on which only around 70 protein coding genes are located (Maan et al. 2017), affects not only the physical properties of the chromatin, but also the regulation of gene expression.

We further explored gene co-expression using weighted correlation network analysis (WGCNA), in order to link LOY in AD and our findings from analyses of DMGs and/or DEGs. We identified multiple modules of co-expressed genes that were significantly up-or down-regulated in LOY and many of the module members were differentially methylated. Among the most upregulated modules, both in monocytes and granulocytes, we found those implicated in immune defense and immune responses (Figure 4). This agrees with the notion that neuroinflammation is the key pathological marker of AD (Kinney et al. 2018; Heppner et al. 2015; Jevtic et al. 2017). The genes contributing to the observed enriched functional categories were, among others, interleukins, or interferon genes. Furthermore, the dysregulated (both transcriptionally and in terms of DNA methylation) members of these immune-related modules were for instance transcription factors (e.g. *ZNF496*), or regulators of epigenetic gene silencing, such as *SIRT7* gene (Figure 4). The sirtuins are implicated in age-related changes in neurodegenerative diseases; hence, they can play an important role in AD (Jęśko et al. 2017). Moreover, it was shown that inhibition of *SIRT7* may be beneficial in AD pathogenesis through the regulation of production of reactive oxygen species (Mizutani et al. 2022).

In conclusion, we provide new evidence suggesting that LOY in immune cells plays a role in the pathogenesis of LOAD in males. Our combined analyses of CpG methylation as well as expression analysis identified new candidate genes and we confirm numerous genes already implicated in the pathogenesis of LOAD. Our results are also well aligned with the hypothesis that age-related dysfunction of the immune system cells is one of the major factors contributing to the development of AD. Loss of a single male chromosome is also clearly reflected as higher level epigenetic changes that show an AD-specific pattern, further contributing as a potential biomarker of the disease.

## MATERIALS AND METHODS

### Study group

Blood samples from patients diagnosed with Alzheimer’s disease (AD) and age-matched controls (above 65 years) were collected from male subjects in Uppsala, Sweden and Kraków, Poland for the purpose of estimating LOY levels, previously described in (Dumanski et al. 2021). The availability of sufficient amount of DNA was used to select a group of 73 individuals for complementing the precious study with DNA methylation analyses. Samples from AD patients and controls were collected during January 2015 to May 2018, at the Geriatric/Memory Clinic, Uppsala Academic Hospital Sweden. Additional samples from AD patients were collected from January 2017 to May 2018 at the Clinic of Internal Diseases and Gerontology of the Jagiellonian University in Kraków, as well as control samples collected from December 2015 to May 2018 from the general population of Kraków. The criteria for recruitment of AD patients were ongoing clinically and radiologically confirmed diagnosis, intermediate or severely advanced disease.

The study was approved by the local research ethics committee in Uppsala, Sweden (Regionala Etikprövningsnämnden i Uppsala (EPN): Dnr 2005-244, Ö48-2005; Dnr 2013/350; Dnr 2015/092; Dnr 2015/458; Dnr 2015/458/2, the latter with update from 2018) and the Bioethical Committee of the Regional Medical Chamber in Kraków, Poland (No. 6/KBL/OIL/2014). All participants or next of kin have given their written informed consent to participate. The study was conducted according to the guidelines of the Declaration of Helsinki.

### Sample preparation

16 ml of blood was collected into two BD Vacutainer® CPT™ Mononuclear Cell Preparation Tubes (BD). Peripheral blood mononuclear cells (PBMCs) were isolated following the manufacturer’s instructions. The PBMCs were then washed with PBS, a portion of PBMCs were aliquoted for scRNA-seq. Additional 16 ml of whole blood were collected into two BD Vacutainer® K2 EDTA tubes (BD). Red blood cells were lysed using 1× BD Pharm Lyse™ lysing solution (BD). Isolated white blood cells (WBCs) were washed with PBS. Targeted cell populations were sorted from the isolated PBMCs and WBCs using FACS as described before (Dumanski et al. 2021). In brief, live cells were sorted based on their FSC and SSC. Monocytes were defined based on their size and as CD14+; granulocytes were defined based on their size and granularity. Cells were sorted to achieve the purity of above 96%. Cell fractions were split for downstream DNA or RNA extractions. Cells sorted for RNA extraction was dissolved in RNAprotect Cell Reagent (Qiagen), all cell fractions were then pelleted and frozen in -70 for further processing.

### Estimation of LOY levels

DNA was extracted and quantified from each isolated cell population following protocols as described before (Dumanski et al. 2021). DNA was genotyped using three different versions of SNP-arrays InfiniumCoreExome-24v1-1, InfiniumOmniExpressExome-8v1-3 and InfiniumQCArray-24v1 (Illumina). All genotyping experiments were performed following the manufacturer’s instructions at the Science for Life technology platform SNP&SEQ at Uppsala University, Sweden. All included experiments passed strict quality control at the genotyping facility. Additional QC criteria as well as calculation of mLRRY as described before (Dumanski et al. 2021). The percentage of cells with LOY (%LOY) in each sample was estimated using a formula described previously, i.e., 100*(1-(2^(2*mLRRY)^)) (Danielsson et al. 2019).

### Bisulfite conversion of DNA samples

Based on the DNA concentrations determined by Quant-iT PicoGreen dsDNA Assay (Thermo Scientific), 250 ng of each DNA sample were used in bisulfite conversion of methylated CpG sites using the EZ DNA MethylationTM Kit (Zymo Research). The bisulfite converted DNA was eluted used for methylation analysis according to the manufacturer’s protocol.

### Methylation analysis

Methylation profiling was performed with the Infinium assay using the MethylationEPIC_v-1-0 array (Illumina) according to the manufacturer’s protocol. The scanning of the EPIC arrays and determination of signal intensities were performed by the iScan System (Illumina). Intensities were normalized using Illumina’s internal normalization probes and algorithms, with background subtraction. The analysis in this project were performed by staff at the SNP&SEQ Technology Platform, Uppsala University, Sweden. The SNP&SEQ Technology Platform is a part of the National Genomics Infrastructure (NGI) and SciLifeLab, Sweden.

### Analysis of DNA methylation data

The raw IDAT files were read into R using the readEPIC function from the wateRmelon package. Next, lumiMetyC (with quantile normalization) and BMIQ functions from the lumi and wateRmelon packages respectively were used to perform data normalization. Next, the data were filtered by minimum detection p-value and all probes overlapping with known SNPs were removed. Probe annotation was performed within R using the package *IlluminaHumanMethylationEPICanno*.*ilm10b4*.*hg19*. Here we focused on several different annotation categories, such as *UCSC_RefGene_NAME, Relation_to_Island* (OpenSea, Island, N_Shore, N_Shelf, S_Shore, S_Shelf), *Regulatory_Feature_Group* (Promoter_Associated, Gene_Associated, NonGene_Associated, Unclassified), *UCSC_RefGene_Group* (TSS1500, TSS200, 5’UTR, 1stExon, Body, ExonBnd, 3’UTR).

To identify statistically significant differences in methylation between the compared groups we used the *dmpFinder* function from the *minfi* package. Probes located on chromosome Y were removed prior to the analysis. Significant DMPs were called when absolute average change in methylation between sample groups was at least 0.5 (M-value) and adjusted P-value<0.05. To intersect DMPs with gene regions, we used the UCSC based annotations and the promoter regions of UCSC genes were defined by a genomic window of +/-2 kb from TSS using the *promoters* function from the GenomicRanges R package (Lawrence et al. 2013). Visualization of genomic neighborhood was performed using *Gviz* package. Gene annotations were plotted for *hg19* version of the human genome using UCSC-based gene annotations.

### Bulk RNA extraction and sequencing

RNA-seq data of sorted cell populations were processed during the previous study (Dumanski et al. 2021). Briefly, cell pellets were used for extraction of RNA using RiboPure™ RNA Purification Kit (Thermo Fisher Scientific), which was further purified from possible DNA contaminations with TURBO DNA-free™ Kit (Thermo Fisher Scientific), and finally followed by quality assessment with Agilent 2100 Bioanalyzer (Agilent Technologies). Libraries of all samples were prepared using the Ion AmpliSeq Human Gene Expression kit (Thermo Fisher Man0010742), loaded on Ion 550 chips, and sequenced on Ion S5 XL system. All sequencing library handling steps were done using manufacturers’ recommendations.

### Bulk RNA-seq data processing

The sequenced AmpliSeq data were basecalled with the Ion Torrent Suite Sever 5.8.0.RC2 software (Thermo Fisher). The generated reads were aligned to the human transcriptome reference (hg19 AmpliSeq Transcriptome ERCC v1) with TMAP mapper. Next, the raw gene expression counts for all amplicon targets in the assay (20,183) were merged to create separate count matrices for each of the two cell types (granulocytes and monocytes). Count data were processed with the R library *edgeR* version 3.28.1 (Robinson et al. 2010). We kept only the genes with expression levels above 1 count per million in at least six samples to remove low quality data. Further assessment of data quality using principal component plots revealed sample grouping corresponding to sequencing batch and patient source. The batch effects were thus adjusted using the *ComBat_seq* function from the *sva* package (version 3.35.2) (Leek & Storey 2007). Genewise Negative Binomial Generalized Linear model was applied to test for differential expression of genes between the AD patients with and without LOY. We considered genes to be significantly differentially expressed by applying a threshold of <0.05 to the corrected p-values (FDR, Benjamini-Hochberg adjustment).

### Building gene regulatory networks with WGCNA

We run the weighted correlation network (WGCNA) analysis in R (Langfelder & Horvath 2008) aiming at reconstruction of gene networks associated with LOY. The analyses were conducted in a step-by-step manner, as explained in the WGCNA R package tutorial, with some small modifications. The normalized gene expression matrices (granulocytes and monocytes) were inspected with the *goodSamplesGenes* function. To calculate the adjacencies, we used the *sigmoidAdjacencyFunction* instead of the standard *adjacency* function for which the *mu* (shift) and *alpha* (slope) parameters were chosen after a series of tests for the most optimal combination and the input matrix of similarities was generated from the expression matrix using the *cor* function. From a given adjacency matrix, we calculated topological overlap matrix and the corresponding dissimilarity, using the *TOMsimilarity*, setting the *TOMtype* to *signed*. Minimum module size was set to 30. This way, we obtained the modules composed of genes that were highly associated with each other. Subsequently, we used *limma’s* functions *lmFit* and *eBayes* to test which modules have the highest fold change in relation to LOY. Lastly, members of each module were labeled to annotate the genes as belonging to any of the following categories (DMP-related, DMR-related, DEG, TF). Members of each module were tested for gene ontology and KEGG pathway enrichment.

### Preparation of samples for single cell RNA sequencing (scRNA-seq)

The aforementioned PBMCs were diluted in 1X PBS with 0.04% BSA to a concentration of 10^6^ cells/ml. The PBMCs were then loaded on a Chromium Single Cell 3′ Chip v2 (10X Genomics) for sequence library preparation according to manufacturer’s protocol. The single cell libraries were sequenced on NovaSeq instrument (Illumina). The single cell library preparation and sequencing were performed at the Science for Life technology platform SNP&SEQ, Uppsala University, Sweden.

### Single cell RNA-seq data processing

scRNA-seq data were obtained from the previous project (Dumanski et al. 2021). Samples matching the ones used in bulk RNA-seq analyses (26 patients, 9 AD *LOY*, 17 AD *non-LOY*) were processed using the standard protocol for the 10X Genomics data. Briefly, the UMI count matrices were obtained using the *CellRanger* v4.0.0 software by mapping against the GRCh38 version of the human genome. The recorded UMIs for each gene in each cell were subsequently processed with the R package *Seurat* v3.2.3 (Stuart et al. 2019). We excluded cells not fulfilling any of the following criteria: at least 400 genes and not more than 3,000 genes per cell, at least UMI 800 and not more than 12,000 UMI per cell, maximum 10% of mitochondrial reads. We further excluded genes that were expressed in less than ten cells. Each separate dataset was log-normalized, scaled and subjected to cluster analyses under various resolutions, as well as cell type labeling using the *SingleR* version 1.06 (Aran et al. 2019) package against the *MonacoImmuneData* provided by the celldex package (version 0.99.1). The latter process was run twice to provide general cell type label (*label*.*main*) and a fine-grained cell type classification (*label*.*fine*). Based on the agreeing cluster/immune label labels we selected only the cells identified as *Monocytes* and performed data integration using standard Seurat protocol designed for the task. Two-dimensional visualization of the scRNA-seq data was done using UMAP and based on the first 30 dimensions of the supervised PCA. Lastly, cells were classified as non-LOY or LOY subject to presence or total lack of detectable expression from chromosome Y genes, respectively.

To test for differentially expressed genes using scRNA-seq we used the *FindAllMarkers* functions in *Seurat*. The default *wilcox* test was applied to identified DEGs based on the following criteria: >0.2 log-fold change, expressed in >=10%, and Bonferroni-corrected p-values <0.05.

### Annotation of genes, gene ontologies and KEGG pathways

Gene set enrichment analyses were performed in R using the *Cluster Profiler* package. Gene symbols (either DMG or DEG) were mapped to Entrez ID. As a background for enrichment analyses for DEGs we used a set of all genes expressed in each tissue (based on bulk RNA-seq data). DMGs were tested against the background of all genes.

### Analyses of TADs and histone modification marks

We use HiC data from the THP-1 monocytic cell line (hic files, https://www.encodeproject.org/experiments/ENCSR748LQF/). TAD boundaries were identified from the HiC data using the arrowhead algorithm (part of the Juicer software, https://github.com/aidenlab/juicer, (Durand et al. 2016). The downstream analyses were performed in R using the Genomic Ranges package (Lawrence et al. 2013). We first merged the adjacent overlapping TADs, capped UCSC genes under each TAD, as well as the individual CpG probes. Differential methylation of TADs was performed using the R package limma (Ritchie et al. 2015).

To analyze histone modification marks in monocytes, we used the RoadMap epigenomics data http://www.roadmapepigenomics.org/data/ from the monocytic CD14 primary cells. Specially, we obtained the BED files from H3K4me3 and H3K27ac histone marks. Manipulation (overlap, merge, intersect) of these coordinate-based data was done in R using the Genomic Ranges package (Lawrence et al. 2013). Active promoters were identified as overlapping regions of H3K4me3 and H3K27ac. Intergenic enhancers were defined as regions of H3K27ac not overlapping H3K4me3. Each of the above-mentioned regions of histone marks had DNA methylation values assigned by capping individual CpG probes.

### Data availability

The DNA methylation data used in this study are available from the authors upon a reasonable request. The bulk and single-cell RNA-seq datasets are available upon a reasonable request from the authors of the original publication (Dumanski et al. 2021). ChipSeq data used to delineate the regions of active promoters and enhancers in CD14 primary cells were retrieved from the Roadmap Epigenomics Project Data stored at GEO https://www.ncbi.nlm.nih.gov/geo/roadmap/epigenomics/.

### Code availability

Unless otherwise stated, all the data processing and visualization of the results was done in R (version 3.6.3). The main code used for processing of single cell data is available under https://github.com/jakalssj3.

## Data Availability

All data produced in the present study are available upon reasonable request to the authors

## Abbreviations

LOY: loss of chromosome Y
AD: Alzheimer’s disease
DMP: differentially methylated probe
DEG: differentially expressed gene
DMG: differentially methylated gene

## ACKNOWLEDGEMENTS

We thank the anonymous patients and volunteer healthy controls for sample contribution and information provided in the questionnaire. We are grateful to Dr. Natalia Filipowicz for coordination of the project, Drs. Eva Tiensuu Janson and Bozena Bruhn-Olszewska for critical review of the manuscript and Drs. Lars A Forsberg and Jonatan Halvardson for access to published RNA-seq data. This study was supported by grants from the Foundation for Polish Science under the International Research Agendas Programme (grant number MAB/2018 /6; co-financed by the European Union under the European Regional Development Fund), Swedish Heart-Lung Foundation (grant number 20210051), the Swedish Research Council (grant number 2020-02010), Swedish Cancer Society, Hjärnfonden, and Alzheimerfonden to J.P.D.

## CONTRIBUTIONS

E.RB., H.D. conducted laboratory experiments;

M.J., J.M. analyzed the data;

J.P.D. conceived of the idea, obtained funding;

J.M., J.P,D. supervised the project;

M.J., J.M., D.S., M.I., J.P.D. contributed to interpretation of the results;

M.J., J.P.D., J.M. drafted the manuscript;

D.S. contributed to data acquisition;

E.RB., H.D., M.S., J.B., K.W., J.J., J.R., V.G., A.KR., L.K. - contributed to selection, collection and preparation of patient samples

All co-authors read and approved the submission.

