## Supplementary material for "Loss of Y is associated with multi-omic changes in immune cells from Alzheimer’s disease patients": Figure legends

**MAIN FIGURES**

**Figure 1: Results from analyses of LOY in selected populations of leukocytes from Alzheimer’s diseases patients and age-matched controls and the distribution of their DNA methylation profiles**.

**A:** General distribution of % of cells with LOY among the sampled cell types. P-values from Mann–Whitney pairwise tests are present above the boxplots.

**B:** Distribution of %LOY in granulocytes and monocytes stratified by the patient cohort. Alzheimer’s disease (AD) patients show the highest span of LOY values with a bimodal distribution as seen from the background violin plots. Red dotted line corresponds to a threshold at which at least 30% of cells have LOY, hence dividing the samples into the LOY and non-LOY groups. P-values from Mann–Whitney (MW) pairwise tests are above the boxplots.

**C:** Distribution of %LOY in granulocytes (X-axis) and monocytes (Y-axis). Every point in the main panel represents a measurement of %LOY for matched samples collected from the same patients. Narrow panels along each of the axes show results for LOY levels for unmatched samples in each cell type. Red dotted line denotes the 30% threshold used to divide the samples into LOY and non-LOY groups.

**D:** Distribution of differences in genome-wide average methylation values among AD patients only. The average values for non-LOY samples were subtracted from the corresponding values from LOY samples (each value represents a single CpG probe on the EPIC array, n=835424). Values around 0 on the X-axis correspond to no difference, values skewed towards the right side denote higher methylation values in the LOY samples. Orange and purple lines denote density of values for granulocytes and monocytes, respectively.

**E:** Distribution of differences in the genome-wide average methylation values among AD patients for a selected subset of CpG probes. Probes within promoters (+/-2 kb from transcription start site of genes, dashed line) show a pronounced bias towards hypermethylation in LOY samples, both in granulocytes and monocytes. Probes annotated as associated with genes (overlapping any of the genes’ structure, dotted line, UCSC-based annotation) show a bias towards hypomethylation in LOY samples. P-values from Mann–Whitney (MW) tests comparing given difference vs. same difference for all probes are shown.

**Figure 2: Results from analyses of significant differential methylation in LOY vs. non-LOY among AD patients.**

**A:** Volcano plots of the genome-wide differential methylation analyses in granulocytes and monocytes. Black points denote all tested CpG sites. Red dots represent the significant DMPs (FDR <0.05, avDiff >0.5). The direction of methylation change (X-axis) is shown in reference to the LOY samples (hypo – less methylated in cells with LOY, hyper – more methylated in cells with LOY). Orange and purple colors denote DMPs that are differentially methylated only between LOY vs. non-LOY samples in AD (hence AD specific). Boxplots on the bottom show the magnitude of methylation change for all sites versus sites that are specific to the AD samples.

**B:** Venn diagram comparing the number of hyper- and hypomethylated DMPs in granulocytes and monocytes identified by comparing LOY to non-LOY samples among AD patients. 67 hyper- and 819 hypomethylated sites are shared between granulocytes (orange) and monocytes (purple).

**Figure 3: Results from differential expression analyses in AD samples and the correspondence of promoter-related methylation with the observed expression patterns.**

**A:** Volcano plots of the genome-wide differential expression analyses using bulk RNA-seq data of AD samples. Red dots represent genes with significant change in expression (DEGs, FDR <0.05). Direction of expression change (X-axis) refers to the LOY samples (up – higher in LOY, down – lower in LOY).

**B:** Venn diagrams comparing gene sets derived from the differential expression and differential methylation analyses. Comparison includes only the DMGs with sites located specifically within genes’ promoter regions. Values in parentheses (grey font) correspond to DMGs that had non-zero expression in the bulk RNA-seq data.

**C and D:** Selected examples of promoter-associated sites with significant change in methylation in granulocytes (C) and monocytes (D), juxtaposed with a change in expression of an associated gene. Boxplots show methylation and expression values in one of the two compared groups – LOY or non-LOY in AD patients. Hypomethylation in LOY is associated with upregulation of the corresponding gene’s expression in LOY. FDR values below probe ID or gene symbol as derived from the differential tests.

**E and F:** Genomic neighborhood of the promoter region of two selected differentially expressed genes from granulocytes (E) and monocytes (F). All genes are upregulated in LOY group in AD patients and have a hypomethylated site within their promoters (as shown in C and D). Top panel (DNA methylation) shows neighboring CpG probes located within the promoter region and their DNA methylation values in LOY (orange) and non-LOY (green) samples in AD. Second and third panels from top show the identified significant differentially methylated sites and regions, respectively. Panel named “UCSC genes” shows annotation of the gene’s structure. Blue rectangles show exons and introns are shown as thin lines. Arrows on introns show the direction of the gene from its 5’ side. TSS denotes site of start of transcription. The bottom-most panel shows CpG islands located in the vicinity of TSS. Dark green denotes CpG islands, and their shores are in light green. Chromosome name and the genomic coordinates are shown above each plot.

**Figure 4: Co-expression analyses identify regulatory gene networks that link expression and methylation support association to LOY in AD**.

**A:** Weighted gene co-expression network (WGCNA) analysis using bulk RNA-seq data from granulocytes and monocytes identified 8 and 11 gene modules, respectively, that were significantly differentially expressed between the LOY and non-LOY groups in AD patients. Boxes contain size of each module, number of member genes that are differentially expressed (DEGs) or differentially methylated (DMGs). Expression change of each module is represented as a scale ranging from red (upregulated in LOY) to blue (downregulated in LOY). Color-based module labels were generated automatically by WGCNA and are shown below the heatmap.

**B: Correspondence of set-specific modules.** Granulocyte modules are shown on the horizontal axis. Monocyte modules are shown on the vertical axis. Numbers within cells in the matrix correspond to the number of genes common between each individual module. Color shading of cells denotes the significance of the correspondence based on Fisher's exact test – darker shading corresponds to higher significance with a scale of -log10 /P-value.

**C:** WGCNA-based “tan” module in monocytes, consisting of 305 genes (heatmap rows), shows expression changes that highly correlated with the LOY status of samples (heatmap columns, p-value 0.0008). Scaled gene expression values shown on the heatmap range from high in LOY (red) to low in LOY (blue). Rows (genes) and samples (columns) were clustered based on their expression values using the hierarchical clustering method. Large portion of the module consists of genes identified as DEGs (n=165, 54%), majority of which are upregulated in LOY. Many of the member genes have an associated significant DMP and/or DMR (n=15) which are mostly hypomethylated. Twenty genes in the module are important transcription factors (TF). Module eigengene (representative of the gene expression profiles in the module) was defined as the first principal component of the expression matrix of the module. Eigengenes expression pattern among all samples is shown below the heatmap.

**D:** Results of gene ontology enrichment analysis of the “lightcyan” and “purple” modules members (166 and 268 genes, respectively) against the background ontology of all genes expressed in granulocytes. There is a significant enrichment (X-axis) of terms associated with immune/defense responses (Y-axis). Size of the circle corresponds to the number of genes representing the given GO category. Color of the circle denotes the adjusted p-value from the enrichment analysis.

**E:** Results of gene ontology enrichment analysis of the “tan” module members (305 genes) against the background ontology of all genes expressed in monocytes. There is a significant enrichment (X-axis) of terms associated with immune/defense responses (Y-axis). Size of the circle corresponds to the number of genes representing the given GO category. Color of the circle denotes the adjusted p-value from the enrichment analysis.

**Figure 5: Single cell RNA-seq data reveal differences in cellular proportions and distinct expression profiles of monocyte DEGs with respect to LOY status of samples and cells.**

**A:** Distribution of cell type frequencies among monocytes as analyzed using single-cell RNA-seq data. Single cells were labelled using the Monaco et. al 2019 immune cells reference, which allowed to identify monocytes and their three main subtypes – classical, non-classical and intermediate monocytes. LOY status, based on the expression of Y-linked genes was inferred for all individual cells. There is a significant difference in the frequencies of the three subtypes between the identified LOY-based cell types.

**B:** Examples of genes identified as DEGs using bulk RNA-seq data in monocytes that are supported by the scRNA-seq data from corresponding patients. Many DEGs from bulk RNA-seq analyses are in fact characterized by distinct expression patterns that derive from their actual subtype.

**Figure 6: LOY involves global higher level epigenetic and regulatory changes**.

**A:** Active promoters, defined as intersecting regions of H3K4me3 and H3K27ac histone marks (+/- 2 kb from gene transcription start site), show predominant hypermethylation in LOY (average DNA methylation value of all probes within histone marks boundaries). This observation holds true both in AD patients and healthy controls. Intergenic enhancers, defined by H3K27ac histone marks without overlap of H3K4me3 and at least 5 kb away from genes’ boundaries, are mostly hypomethylated in LOY in AD, while healthy controls peak close to zero imposing prevalent lack of significant differences in average methylation values in this cohort.

**B:** Density plot showing methylation landscape of LOY and non-LOY samples bound by the topologically associated domains (TADs) from THP-1 monocytic cells. Each TAD is represented by an averaged sample-specific methylation value of all probes falling within its genomic boundaries. Lines correspond to the change in average methylation value between LOY and non-LOY samples in healthy controls (black) and AD patients (red). There is a significant difference between the two cohorts (Mann-Whitney test) and AD patients show a strong shift towards hypomethylation in LOY, which such big shift is not observed in healthy controls.

**SUPPLEMENTARY FIGURES**

**Supplementary Figure S1**: **Distribution of differences in genome-wide average methylation values including all sampled groups.**

Groups compared included LOY controls, non-LOY controls, LOY AD and non-LOY AD patients. The average values per CpG site were subtracted from each other between two sample groups. Values around 0 on the X-axis correspond to no difference, values skewed towards the right side denote higher methylation. Each panel is named after the type of CpG probes, as annotated in the Infinium MethylationEPIC Manifest file. Number of probes in each category is provided above each panel. Numeric values plotted next to the density curves show median value in average methylation difference per comparison.

**Supplementary Figure S2**: **Genomic distribution of the identified hyper- and hypomethylated DMPs as compared to the distribution of all probes on the EPIC chip.**

**A.** Distribution of probes and DMPs in relation to gene annotations (UCSC_RefGene_Group column in the Infinium MethylationEPIC Manifest). The upper panel corresponds to granulocytes, the bottom panel shows monocytes. P-values shown on the panels were derived from hypergeometric tests of proportion of significant sites vs. all sites in each category.

**B.** Distribution of probes and DMPs in relation to CpG islands (Relation_to_Island column in the Infinium MethylationEPIC Manifest). The upper panel corresponds to granulocytes, the bottom panel shows monocytes. P-values shown on the panels were derived from hypergeometric tests of proportion of significant sites vs. all sites in each category.

**Supplementary Figure S3**: **Distribution of the identified differentially methylated sites and associated genes.**

**A&B:** Manhattan plots of distribution of the identified differentially methylated sites from granulocytes and monocytes. Colored points correspond to significant DMPs (FDR <0.05, average difference >0.5), the remaining grey dots are all other tested sites. Values on the Y-axis are -log10 of FDR. Selected DMP-related genes, i.e., genes with at least one differentially methylated probe, are shown along the x-axis above for hyper- and below for hypomethylated probes. Genes were selected based on having a significant DMPs that is located within promoter region (+/- 2kb from TSS) and is among the sites with the lowest LOY-associated FDR value.

**C:** Venn diagram showing intersection between the identified differentially methylated genes (DMGs) in granulocytes and monocytes. DMGs have at least a single differentially methylated probe located anywhere in the gene’s boundary.

**D:** Venn diagram showing intersection between the identified differentially methylated genes (DMGs) in granulocytes and monocytes and the LATE (LOY-associated transcriptional effect) genes as identified in Dumanski et al. 2021.

**Supplementary Figure S4**: **Comparison between DEG and DMG sets in granulocytes and monocytes.**

**A.** Venn diagram showing comparison between up- and downregulated differentially expressed genes found by comparing LOY to non-LOY samples in AD in granulocytes and monocytes.

**B.** Venn diagram showing comparison between DEGs and DMGs identified in granulocytes and monocytes.

**Supplementary Figure S5: Gene ontology enrichment analyses of additional WGCNA-based gene modules from granulocytes.**

**A.** Results of gene ontology enrichment analysis of the “blue” module members (954 genes) against the background ontology of all genes expressed in granulocytes. This module was found to be upregulated in AD LOY samples. There is a significant enrichment (X-axis) of terms associated with viral transcription and gene expression, as well as neutrophil degranulation and activation in immune responses (Y-axis). Size of the circle corresponds to the number of genes representing the given GO category. Color of the circle denotes the adjusted p-value from the enrichment analysis.

**B.** Results of gene ontology enrichment analysis of the “green” module members (367 genes) against the background ontology of all genes expressed in granulocytes. This module was found to be downregulated in AD LOY samples. There is a significant enrichment (X-axis) of terms associated with protein heterodimerization or nucleosome and chromatin assembly and DNA packaging (Y-axis). Size of the circle corresponds to the number of genes representing the given GO category. Color of the circle denotes the adjusted p-value from the enrichment analysis.
